# Modeling the drivers of differential Typhoid Conjugate Vaccine (TCV) impact in Pakistan: force of infection and age-specific duration of protection

**DOI:** 10.1101/2024.08.30.24312839

**Authors:** Alicia N.M. Kraay, Mohammad T. Yousafzai, Sonia Qureshi, Jillian Gauld, Farah N. Qamar

**Affiliations:** Institute for Disease Modeling, Bill & Melinda Gates Foundation, Seattle, WA; Department of Paediatrics & Child Health, The Aga Khan University Hospital, Stadium Road, P.O Box 3500, Karachi 74800, Pakistan

## Abstract

**Background:** While trials have demonstrated high efficacy of typhoid conjugate vaccine (TCV), data on effectiveness are limited. We report initial impacts and predict future benefits of TCV from two provinces in Pakistan.

**Methods:** We used blood culture-confirmed typhoid cases from the Surveillance for Enteric Fever in Asia Project (SEAP) and Impact assessment of Typhoid conjugate vaccine following introduction in Routine Immunization Program of Pakistan (ITRIPP) to estimate the population-level impact of vaccination (2018-2023). We used regression models to estimate initial impacts and an agent-based model to predict future benefits.

**Findings:** In Sindh, typhoid incidence was higher and cases were younger compared with Punjab. TCV reduced incidence by 48.9% in Sindh (95% CI: 47.3-50.3%) and 66.2% in Punjab (95% CI: 64.7%, 67.6%) over the first 2 years after vaccine rollout but declined each year. In Sindh, waning was quicker and models predicted that population incidence would stabilize near pre-vaccine levels in 2024. An additional campaign could provide short-term, but not long-term, benefits. In contrast, in Punjab, incidence is projected to remain low for several years, and the standard vaccine program may be sufficient. However, follow up data from Punjab are needed to better characterize waning immunity.

**Interpretation:** TCV has reduced incidence in Pakistan, but protection varies by site. Standard vaccine policy may be sufficient to control incidence in settings with moderate transmission. However, in settings with particularly high incidence and/or short duration of protection, alternative strategies to reduce the force of infection may be needed.

**Funding:** Bill & Melinda Gates Foundation

## Background

Typhoid is an enteric infection with high disease burden in South Asia. In 2017, there were an estimated 14.3 million infections and 136,000 deaths, with the vast majority of these deaths occurring among young children in South Asia (1). While incidence has been declining in many countries, Pakistan continues to suffer from large outbreaks due to a variety of factors, including poor access to adequate water, sanitation, and hygiene infrastructure and increasing prevalence of antibiotic resistance, which makes treating cases more difficult (2). Indeed, Pakistan is now considered to be the main global source of antibiotic resistant cases (3), with 70% of cases being extensively drug resistant in 2020 (4).

One tool to reduce typhoid is the typhoid conjugate vaccine. In clinical trials, TCV reduced blood confirmed typhoid incidence by 79% in Nepal (5) and 81% in Malawi (6). These initial trial data were published on the basis of 2 years of follow-up data and formed the evidence base that led to TCV approval and endorsement by the WHO (7). In Pakistan, a surveillance study found strong efficacy of up to 97% in an outbreak setting (8,9), suggesting similar efficacy was also achievable in Pakistan. More recently, data on individual-level protection after 4 years has been published from Malawi, with a recent paper showing no waning by 4 years after vaccination (6,10). However, longer term population level field data on vaccine impacts after large scale rollout are needed to understand differences between vaccine efficacy and real world effectiveness.

For many vaccines, obtained protection and waning of that protection is known to vary by site (11). One reason for this heterogeneity includes differences in initial vaccine response (12,13), which may interact with the age distribution of cases. Some studies have shown that immunogenicity for typhoid vaccination is age-specific, with older ages showing a potentially stronger response (14). In settings with a younger age distribution (resulting from high transmission intensity), the overall population averaged protection might be lower due to interactions between vaccine response and age-specific incidence. High transmission intensity can also overwhelm expected protection due to high frequency and/or level of exposure. More specifically, the ultimate difference in cumulative risk over follow up narrows over time even if immunological waning does not change in high transmission settings. Given the wide variation in incidence across countries targeted for TCV vaccination (15), studies are needed to assess duration of protection at the population level across sites.

In this study, we show population level impacts of typhoid vaccination for Sindh and Punjab provinces in Pakistan covering 4 years of vaccine rollout in Sindh and 3 years in Punjab. We provide estimates by age group and over time to show the dynamics of protection and differences by site. We then use a transmission model to explore how incidence is expected to change in the future and how vaccine strategy could shape these patterns.

## Methods

### Data

Data are from the laboratory network in Sindh and Punjab provinces from the Surveillance for Enteric Fever in Asia (SEAP) study (15) and ITRIPP (unpublished) include the number of blood confirmed typhoid cases by month and age group (<2, 2-4, 5-9, 10-14, and ≥15 years). q. Based on a comparison of incidence and test positivity over the surveillance period (see SI text and Figure S1), we focus on data from 2019-2023 for calculating impacts in the main text for both provinces. We also considered running Sindh with 2018-2023 data to allow for two years of pre-vaccine data as a sensitivity analysis, but focus on 2019-2023 in the main text to match Punjab.

### Statistical analysis

#### Pre-post Poisson regression model

We used Poisson regression models to estimate the impact of vaccination comparing the pre-vaccine era (reference) with the post-vaccine era. For both provinces, we excluded data collected from February-June 2020 during a national COVID lockdown. For Sindh, we used data from January 2019-November 2019 as the pre-vaccine period and January 2020 and July 2020-December 2023 as the post-vaccine period. In addition to the lockdown months, we excluded the month of December 2019 because the campaign was being conducted during that month, and seroconversion to vaccination takes about 28 days (16). For Punjab, we used January 2019-January 2021 (excluding lockdown months) as the reference and March 2021-December 2023 as the vaccine period, excluding February 2021, the month of vaccine rollout and the vaccine campaign in Punjab.

Population size was included as an offset for all models, based on estimates from the Pakistan Bureau of Statistics from the 2017 census for the urban areas of each province (11). We assumed population growth rates were applied evenly to all age groups. We ran these models for the entire follow up period and all age groups combined, but also ran separate models for each year and age group to better characterize differences in impact by age group and over time. All Poisson regression models were conducted in R (version 4.3.1).

### Simulation model

We used a previously published individual-based typhoid transmission model built in EMOD 2.11 (17). Briefly, susceptible individuals can become infected through transmission through environmental reservoirs. We also considered fitting a direct contact route, but the two modes of transmission in the model were not identifiable and caused difficulties comparing transmission across the two sites. Once infected, the infection is initially latent after which it progresses to acute or subclinical infection. Both acute and subclinical infections can become chronic carriers and may continue to transmit at a reduced rate. We also track the number of infections each agent has experienced with repeated infections being less common (Table S1, Table S2).

The model uses a baseline number of agents of 10,000, with growth rates set to match the regression model. Unlike the regression model, population growth is assumed to be resultant from births outweighing deaths. We simulated this model from 1990 to 2040, with 1990-2015 being treated as a burn in period.

Vaccination is implemented in December 2019 for Sindh and February 2021 for Punjab and is modeled as reducing the risk of infection by an amount proportional to the calibrated vaccine efficacy (see SI for more details). For each province, vaccine introduction is implemented as a one-time campaign among children 9 months to 15 years of age followed by continued routine immunization for 9 month olds. Incidence during lockdown months was excluded from both models for calibration.

### Model calibration and validation

The model was calibrated to the age distribution pre-vaccine as well as overall incidence for each province during the reference and vaccine period (2018-2023 for Sindh and 2019-2023 for Punjab) using a gradient ascent algorithm. Model parameters are shown in tables S1 and S2 and more details of the calibration are shown in the SI.

To validate model predictions, we compared incidence from vaccine simulations with a counterfactual, no vaccine simulation designed to predict what would have happened if the vaccine had never been implemented. We ran 100 stochastic simulations for each parameter set/scenario to capture uncertainty inherent in the transmission process and population dynamics. To facilitate comparison of transmission intensity between the two provinces, we also computed the average age of first infections in 2015-2019, when the model had reached equilibrium, across the 100 stochastic no-vaccination simulations. The age distribution is expected to at its peak prior to vaccination, and inversely related to the force of infection (18).

### Forward simulations

To explore how transmission dynamics might depend on future vaccine strategies we implemented a booster campaign in June 2024, aligning with action considered by GAVI to control the ongoing disease outbreak. We considered: a control scenario with no additional vaccination, a default campaign using the same strategy as the initial campaign, an expanded age campaign, where adults over 15 were vaccinated, and an additional school age dose. We also explored whether combining these interventions led to additional impacts.

## Results

In the population overall, reported cases in Sindh were roughly 4x as high as in Punjab pre-vaccine, with a younger age distribution (Figures 1-3). In Sindh, incidence was highest in children under 2, but was also high among 2-4 and 5-9 year olds. Incidence among ≥15 year olds was low. In contrast, in Punjab, incidence was lowest in children under 2 years of age and peaked among 5-9 year olds, with over 30% of cases 15 years of age and older.

**Figure 1.**
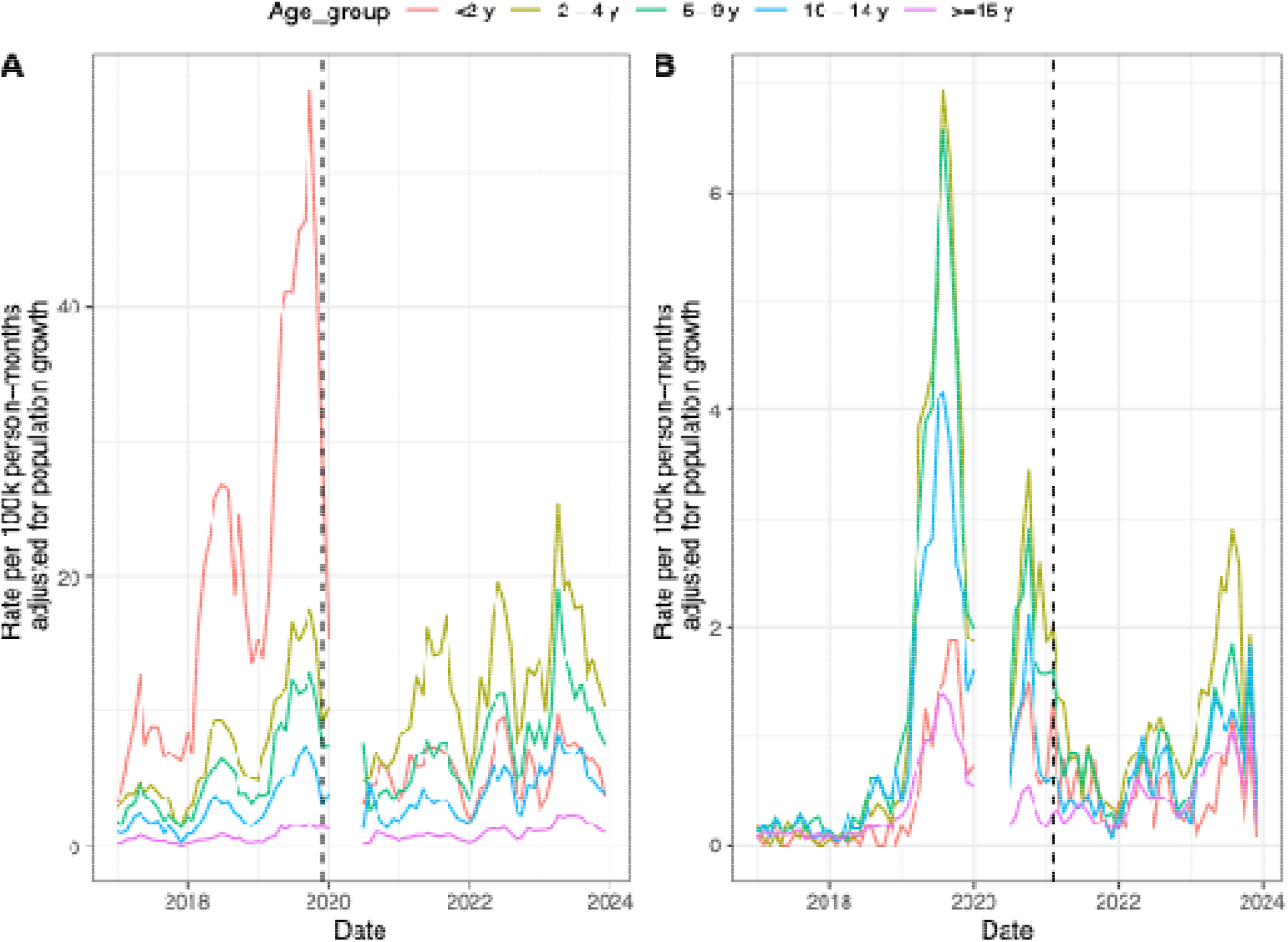
Time series of blood confirmed typhoid cases over time by age for A) Sindh and B) Punjab. The months of February 2020-June 2020 are excluded from the time series due to a national lockdown that occurred in response to surging cases of COVID-19. Timing of vaccine campaigns in each province are shown with the dotted lines. See Figure S1 for pooled time series collapsed across all ages.

For both provinces, incidence declined after vaccination. However, in Sindh, incidence began to rebound 1.5 years later, largely driven by increased incidence in 2-4 year olds. In Punjab, incidence did not begin to rebound until 2.5 years after vaccine introduction, and the resultant peak was not as pronounced as in Sindh (Figure 1). While the rebound in Punjab was highest among the 2-4 year olds, the overall reduction in vaccine effectiveness was greatest among the oldest age group (≥15 years) (Figure 2).

**Figure 2.**
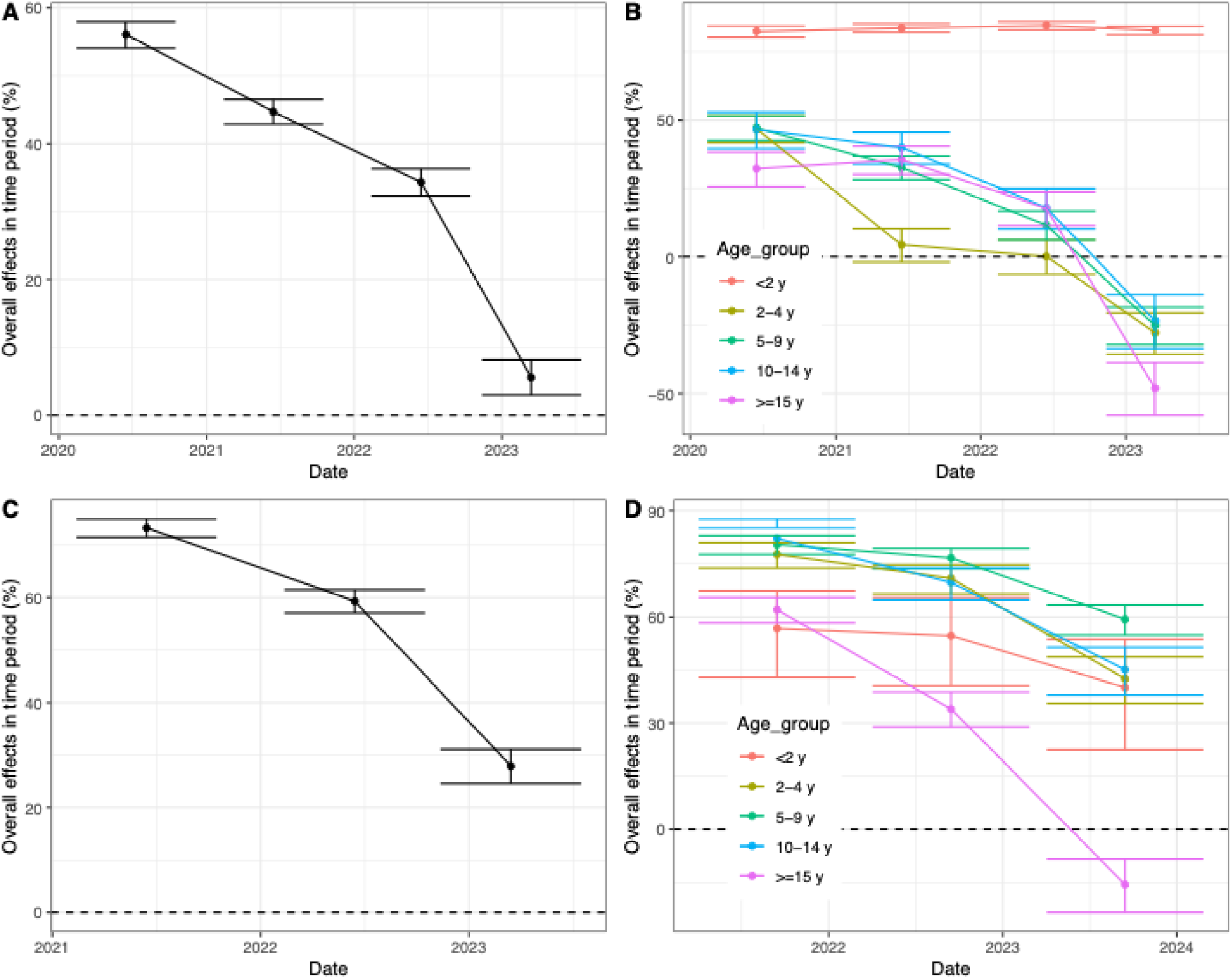
Poisson regression derived vaccine impacts for A) Sindh, all ages (2019 reference), B) Sindh, stratified by age (2019 reference), C) Punjab, all ages, D) Punjab, stratified by age. Note that y-axes are different for each panel. A version with the same y-axis for panels A and C and panels B and D is shown in Figure S3.

The regression models revealed that the pattern in protection was heterogeneous by province, age, and time. For both provinces, efficacy was highest in the first year after vaccine introduction, with an initial estimate of 56.1% (95% CI: 54.1-57.9%) in Sindh and 73.3% (95% CI: 71.5, 74.9%) in Punjab (Figure 2). During the second year after introduction (January-December 2022 in Sindh and March-December 2023 in Punjab), estimates of vaccine impact had fallen in both provinces, with a VE of 44.7% (95% CI: 42.9-46.5%) in Sindh and 59.3% (95% CI: 57.1-61.4%) in Punjab. Thus, average protection over the first 2 years after rollout was 48.9% in Sindh (95% CI: 47.3-50.3%) and 66.2% in Punjab (95% CI: 64.7-67.6%). In Sindh, protection remained high for the youngest age group for all four years, with stable protection of about 70%. However, older age groups appeared to have a decline in protection. For Punjab, protection was most stable for all but the ≥15 age group, who experienced a relatively large decline in efficacy in 2023. While protection was moderate for children <2 years in Punjab, it was lower than in Sindh, with an average protection of 40-60% and a stronger decay over time. Both provinces experienced a large spike in cases in 2023 and had lower vaccine efficacy in that year. If 2018 was included in the reference period for Sindh, the estimates of vaccine efficacy were lower, but the trend over time was similar (Figure S2).

The transmission model for both provinces fit the age distribution pre-vaccine well (Figure 3A, Figure 3B). In Punjab, the model tended to overestimate the number of cases among children 10-14 years of age, but all other age groups were consistent between the model and the data. The model also performed well in capturing the decline and then rebound in incidence after vaccine rollout (Figure 3C, Figure 3D). Fitted parameters suggested longer duration of protection in Punjab compared with Sindh. In Sindh, the estimated duration of immunity among children was much lower, with half of all vaccinated children being fully susceptible before age 5. In contrast, in Punjab, half of young children were expected to be fully susceptible by age 10. Fitted infectiousness was also over four times as high in Sindh compared with Punjab, consistent with higher baseline transmission intensity (Table S2).

**Figure 3.**
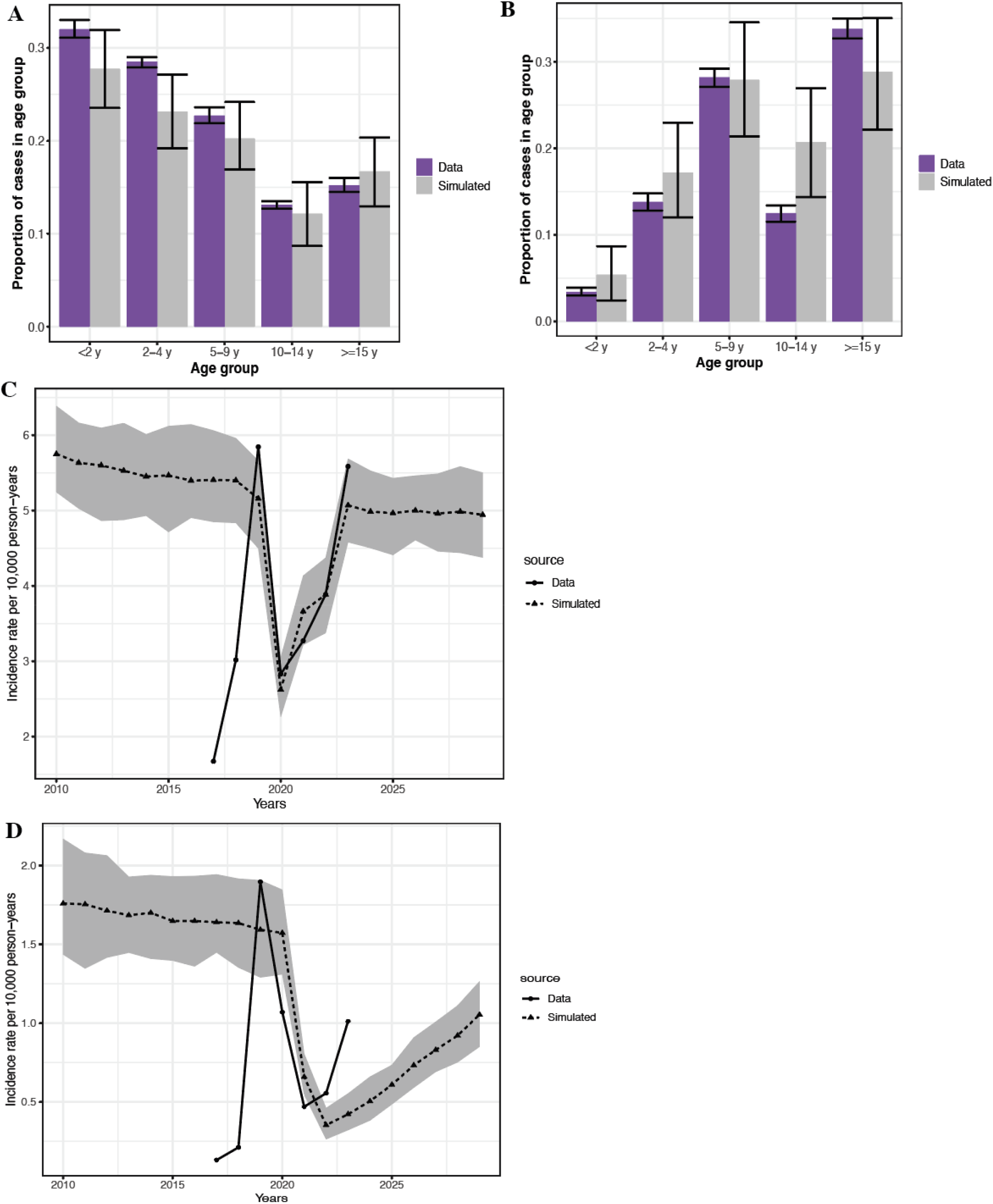
Model calibration figure. A) Simulated and measured age distribution of cases in Sindh pre vaccination and B) Simulated and measured age distribution of cases in Punjab pre vaccination. Pooled time series of blood confirmed reported typhoid cases from data (solid line) vs. model (dashed line and grey ribbon for C) Sindh and D) Punjab. Sindh panels show results with the 2019 reference period.

In Punjab, incidence began to fall late in 2020 after the COVID-19 lockdown but before the vaccine campaign began, which was not captured in our model. The vaccine efficacy obtained from the model closely matched what was seen in the surveillance data when averaged over the first 2 years after vaccine introduction. For Sindh, initial estimates of VE were not as closely matched to the data, but the trajectory was better whereas for Punjab the initial VE estimates matched closely but the rate of rebound was slightly offset. Specifically, for Punjab modeled VE was 68.9% (63.6-73.7%) compared with 66.2% (95% CI: 64.7-67.6%) in the data for 2021-2022. For Sindh, modeled VE was 41.2% (95% CI: 34.3-46.7%) over the first two years compared with 48.9% (95% CI: 47.3-50.3%) in the data for 2020-2021. The differences in rebound of population incidence were strongly patterned by age, with modeled protection being more stable in <2 year olds in Sindh and showing a much faster rebound in older age groups, similar to the primary data. Fits to age-specific incidence are shown in Figures S4-S5.

In the absence of any campaigns or shift in vaccine strategy, incidence was predicted to rebound in both provinces, with incidence being about 25% below pre-vaccine levels in Punjab by 2030 or near pre-vaccine levels in Sindh (Figure 4). For Punjab, the decline in vaccine efficacy was expected to be slower and smaller. In both provinces, a 2024 campaign conferred a substantial short-term benefit. However, in Sindh, this benefit rapidly declined and was followed by a rebound in 2025-2027. In contrast, in Punjab, the rebound in incidence after the campaign was smaller, but no long-term benefit was expected. Augmenting planned campaigns with an expanded age range and/or adding a routine school-age dose did not influence this pattern in Sindh. In Punjab, a school age dose had a greater benefit, appearing to slow the rate of rebound in incidence. Thus, repeated campaigns with similar reach would likely be needed to sustain population level benefits in Sindh. While augmenting the base schedule in Punjab could be helpful, the timeline for such interventions could be slower and could be developed as more data on long-term protection in the province become available.

**Figure 4.**
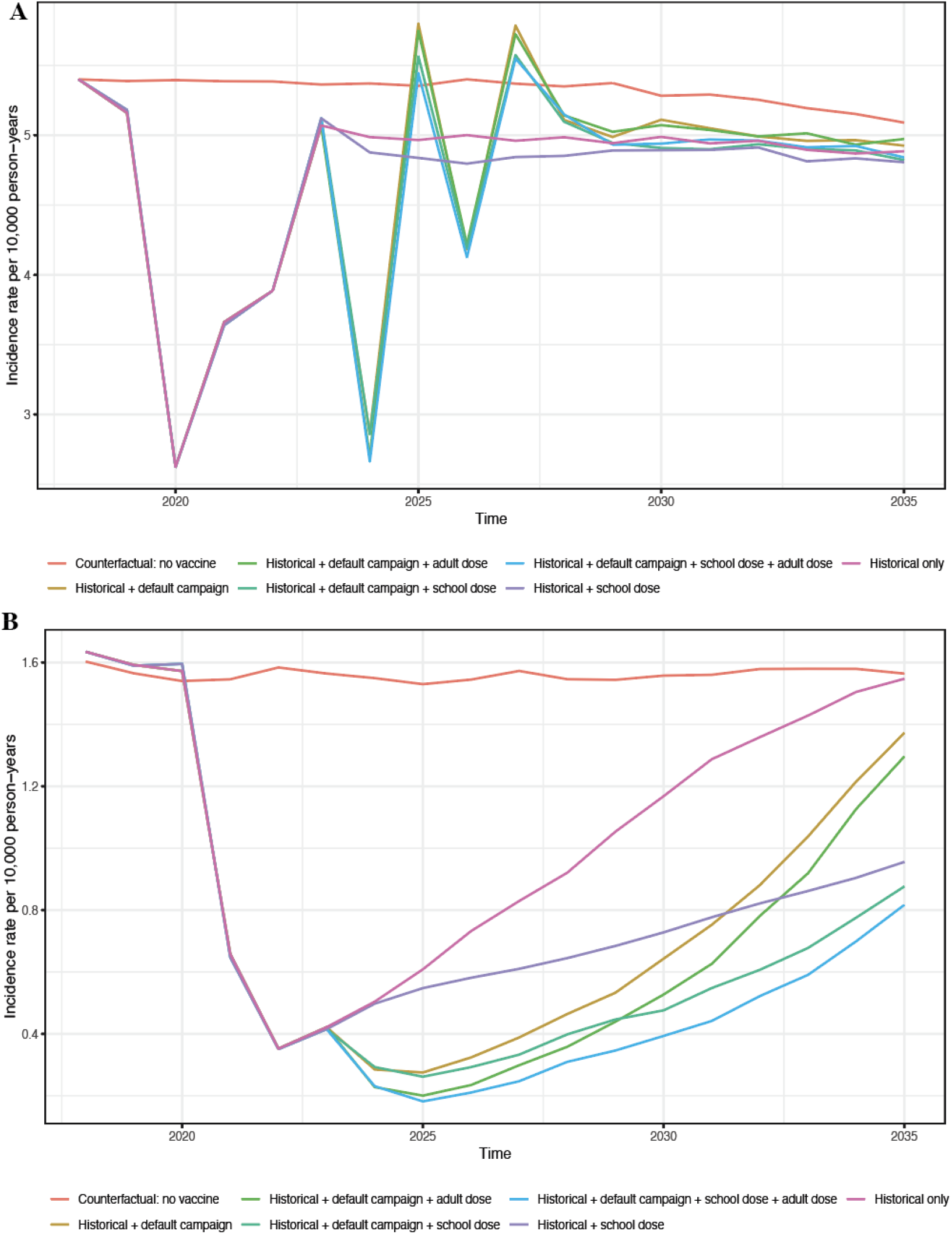
Impact of future vaccine strategies on incidence of blood-confirmed typhoid in A) Sindh and B) Punjab.

As fitted initial campaign coverage was lower in Sindh than Punjab, we also conducted a sensitivity analysis where we simulated the potential impact of vaccination if the follow-up campaign in Sindh were able to achieve 100% coverage, similar to the fitted initial campaign coverage in Punjab. In those simulations, the initial benefit of the campaign was greater (bringing incidence to as low as 2.1 per 10,000 person-years compared with 2.9 per 10,000 for the simulations at the original coverage level), but there was no long-term benefit (Figure S6).

## Discussion

Overall, TCV has substantially reduced typhoid incidence in Pakistan. We found strong impacts shortly after vaccination, particularly for young children. However, protection varied by province and over time. The status quo vaccine policy of routine immunization at 9 months of age may be sufficient to control incidence in settings with moderate transmission intensity and/or slow clinical waning like Punjab. However, in settings with particularly high incidence and/or faster clinical waning, such as Sindh, alternative strategies to reduce the force of infection and tailor vaccine campaigns may be needed. Moreover, complementary strategies will likely be needed to eliminate typhoid.

The differences in overall clinical effectiveness and rebound rates over time were primarily driven by the lower fitted duration of immunity and secondarily by higher transmission intensity in Sindh compared with Punjab. Unlike the other age groups, protection for the youngest group was slightly lower in Punjab compared with Sindh. Effectiveness estimates may have been limited by the extremely low baseline incidence in children <2 years, which reached zero in some of the pre-vaccine years. In Sindh, the estimated duration of immunity among children was much lower. For older age groups, the discrepancy was even greater. All differences in immunity are based on observed clinical protection may be due to multiple factors, including the interplay between antibody decline and force of infection, potential differences in circulating strain diversity, or other unidentified factors. Given that the dynamics of antibody decline have been shown to have similar patterns across countries, even where there are large differences in incidence (19) and appear to be similar in Pakistan (20), it is unlikely that faster antibody titer waning in Pakistan can explain our findings. However, higher antibody titer may be needed to confer protection in high force of infection sites compared with lower transmission intensity settings.

Based on our fitted parameters and the age distribution of infections, the expected force of infection was higher in Sindh compared with Punjab, which may partially contribute to these trends (R_0_ values of 8.0 and 5.6). This transmission intensity for Sindh is higher than has previously been estimated for any location. Bangladesh has roughly been estimated to have an R_0_ of 7, with India having a much lower estimate of 2.8 (21,22). This high transmission intensity may represent a break point above which additional interventions are needed.

While the faster decline in initial benefits in Sindh may also reflect the lower fitted coverage of the initial campaign (Table S2), these differences cannot explain the gap in long term protection (Figure S5). For all forward simulations, we assumed that coverage would likely be similar to the initial campaign to be conservative. However, our sensitivity analysis suggests that a campaign in Sindh with broader reach would likely have a stronger initial impact, but would not impact long term trends. Rather than uniformly increasing coverage, future campaigns targeting high risk subgroups who are commonly infected and more likely to spread disease, such as food handlers, might be beneficial (23).

While our simulations suggest the potential for continued waning in Punjab and thus the possible need for follow up campaigns and/or additional doses in the future, we note that the parameters related to waning immunity in Punjab were particularly uncertain, given only ∼2.5 years of post-vaccination data in Punjab compared with 4 years in Sindh. Thus, additional data in Punjab would be useful to determine if population level protection will continue to decline or will stabilize. These results have implications for future vaccine policy recommendations, which are being considered by WHO to better control typhoid in endemic settings based on data from large-scale rollout that is now becoming available (24).

While beyond the scope of this paper, we note that alternative strategies beyond vaccination may also be able to reduce transmission intensity and will likely be necessary to eliminate typhoid as a public health problem in Pakistan. For example, increasing access to safe water and improved sanitation may also be helpful, particularly for reducing long-cycle transmission through environmental sources (25,26). Treating chronic carriers might also be useful (25). Our results suggest that elimination is unlikely to occur with vaccine interventions alone, similar to the findings of other modeling studies (27).

Our study has a number of limitations. First, we note that surveillance sites were initially limited, particularly in Punjab, and we were thus limited to focusing on 6 years of data in Sindh and 5 years in Punjab, which only included 2 years of pre-vaccine data for each province. The limited length of pre-vaccine data made our comparisons of the pre and post-vaccine periods more sensitive to stochastic effects of high transmission years shortly before vaccination. In 2019, incidence was exceptionally high throughout Pakistan. If incidence expected in the absence of vaccination is substantially lower than occurred in 2019, we may have overestimated the efficacy of vaccination. As other countries prepare to launch their own vaccine campaigns, careful quantification of the baseline burden is important to understand the ultimate impact of vaccination.

From 2020 onwards, our estimates may have also been impacted by external events. At the end of the time series (in 2022 and 2023) there was a large flood coincident with the surge in incidence in both provinces (28). While vaccination did not prevent this large resurgence, this may not be reflective of what we can expect from vaccine impacts going forward. Nevertheless, large storms are expected to become more frequent globally and in Pakistan as climate change progresses (28), so understanding the impacts of vaccination in the context of extreme weather events is important.

Impacts may also have been influenced by the COVID-19 outbreak. COVID-19 may have reduced reported incidence due to reduced social contact, reducing eating out (29), and by reducing health seeking behavior/reporting (30). Conversely, some of the decline in incidence that occurred in 2020 might also be a result of this prior spike in cases, as immunity accrued during 2019 might have reduced incidence the following year. While we removed the lockdown months from model calibration and from the regression models, we cannot completely exclude the impact of COVID-19 on our impact estimates, as the resulting impact on population level immunity may take years to stabilize and this bias may differentially bias impact estimates by province. In Sindh, decline in incidence in 2020 that was unrelated to vaccination may have overestimated estimates of vaccine impact. In Punjab, this bias may have led us to underestimate initial impact estimates, as 2020 was included in the pre-vaccine period. In Punjab, incidence in the latter half of 2020 was substantially lower than in prior years, even prior to vaccine introduction. These patterns may also have been influenced by indirect benefits from the vaccine campaign launched in Sindh several months prior, which we could not capture without a detailed meta-population model.

In this study, we provide population-level estimates of effectiveness of typhoid vaccination in Pakistan. Our results suggest that TCV can provide substantial benefits, but that long-term benefits may be context-specific and require additional efforts to sustain in high burden settings. As more data become available, the expected impacts of typhoid vaccination may become clearer.

## Data Availability

All data in the present study are privileged, but interested researchers may contact the authors to discuss collaboration opportunities.

