## Supplemental Information for "Modeling the drivers of differential Typhoid Conjugate Vaccine (TCV) impact in Pakistan: force of infection and age-specific duration of protection"

**Supplementary Information**

**Reference period selection.** While the SEAP study began in 2016, data collection efforts were more limited earlier in the study, particularly in Punjab, and labs were added throughout the follow-up period. To determine the appropriate reference period for each province, we compared incidence of blood culture confirmed typhoid cases with test positivity over the same period. In general, incidence and test positivity tracked together. However, test positivity began to increase rapidly in Punjab beginning in 2019. Positivity in Sindh appeared to be higher than would have been expected given incidence in 2017 and 2018, suggesting that surveillance was not initially strong enough to capture most cases (Figure S1).

**Figure S1.** Time series of blood culture confirmed typhoid cases over time and culture positivity for A) Sindh and B) Punjab. Incidence is shown in blue and percent positivity is shown in red. Timing of vaccine campaigns in each province are shown by dashed lines.

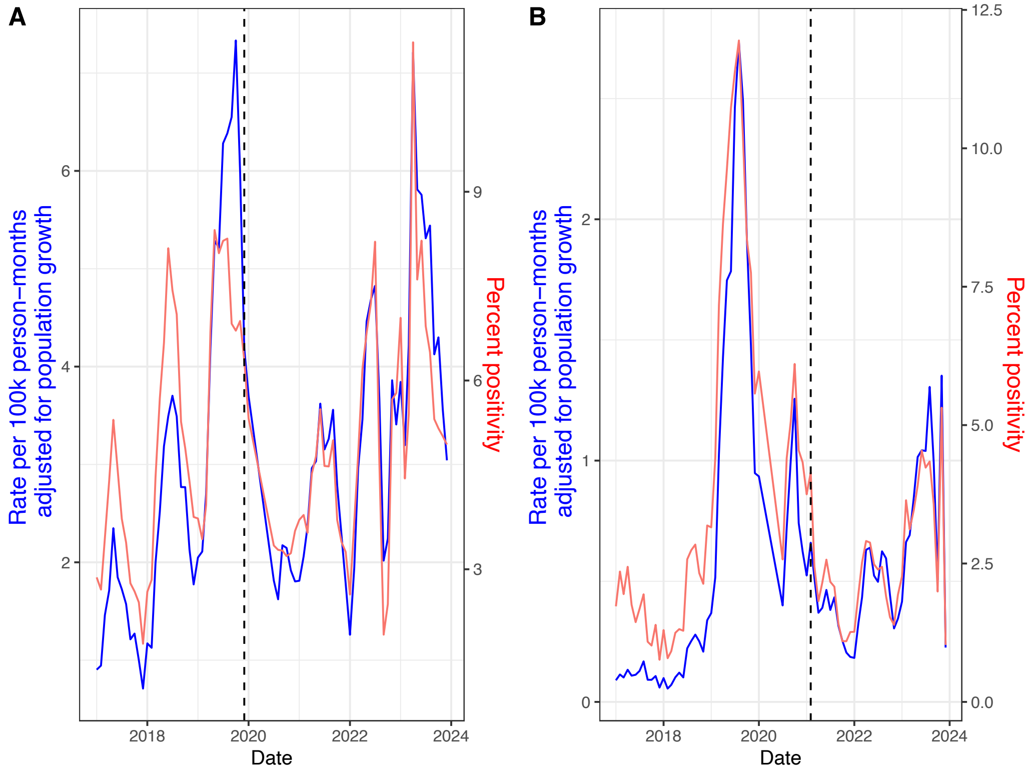

**Figure S2.** Comparison of estimates of vaccine efficacy for Sindh by reference period used. Panels show VE with 2018 included for the full population (A), by age group (B) and the same two panels with only 2019 included as the reference period for the full population (C) and by age group (D).

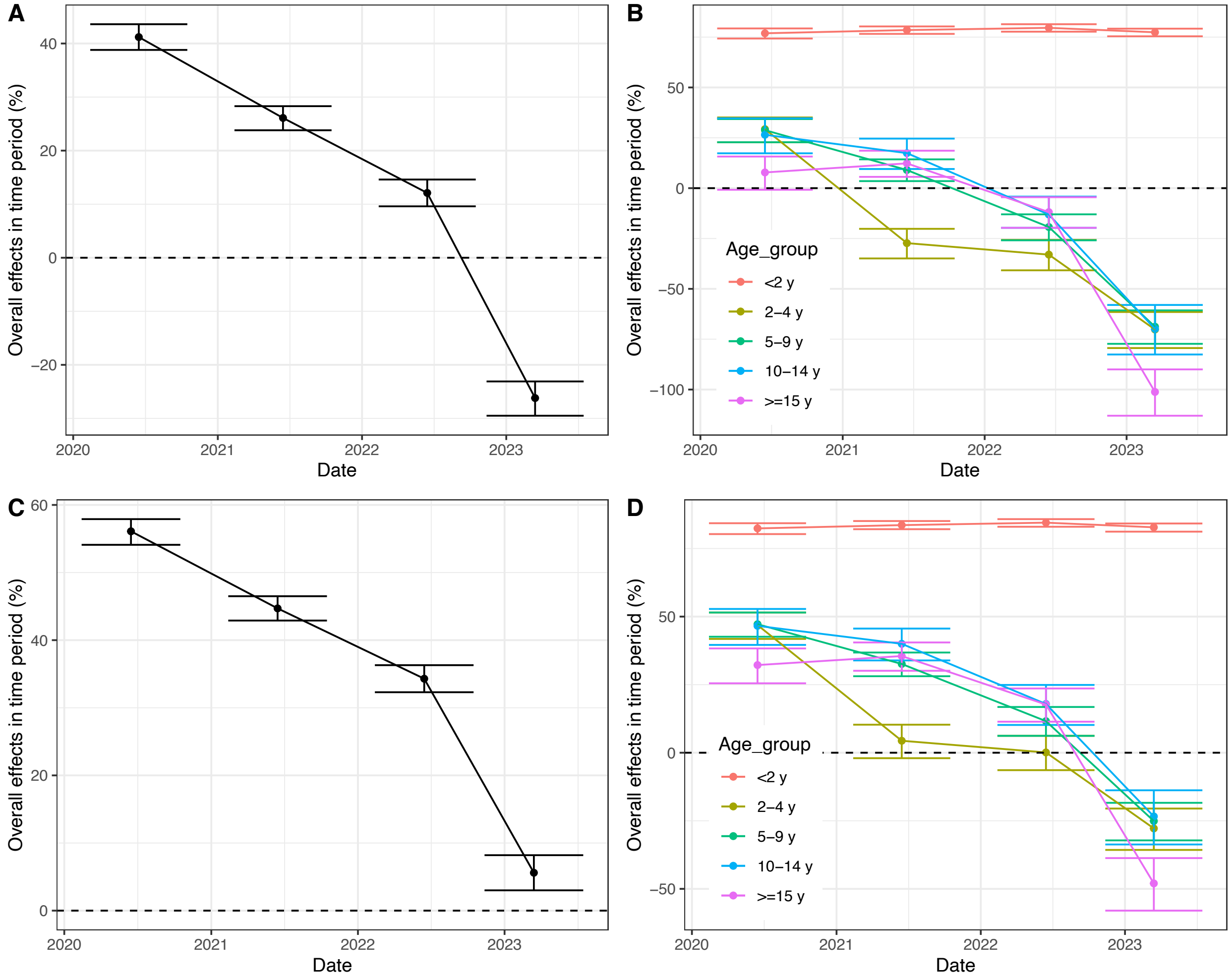


**Calibration details.** We calibrated the model in three steps. In step 1, we calibrated the model to the age distribution of blood culture confirmed typhoid cases prior to vaccine introduction, focusing on January 2019-December 2020 for Punjab and January 2018-December 2019 for Sindh. In step 1 we calibrated the parameters related to the baseline force of infection (infectiousness, lambda).

In step 2, we jointly calibrated parameters related to vaccination (including efficacy, coverage of routine immunization, campaign coverage, and duration of protection from vaccination for each age group) as well as a reporting rate to the age-specific incidence time series for each age group (<2, 2-4, 5-9, 10-14, and 15+) for the full reference period (January 2019-December 2023). Duration of protection was modeled as following a box exponential structure, with an initial period of high protection followed by gradually waning protection given by an exponential decay based on prior literature showing similar patterns for other pathogens (1) and the initially high protection observed before waning across other clinical trials for the typhoid conjugate vaccine (2,3). Because waning protection from the statistical analysis appeared to have distinct patterns for <2 year olds, 2-4 year olds, and 5+ year olds for Sindh, we used these three age groups to fit duration of protection, giving 6 vaccine waning parameters. In Punjab, patterns varied with faster apparent waning among the 15+ age group, so we used <2, 2-12, and 12+ years to define the waning immunity bins to more closely reproduce the decline in clinical protection.

For both provinces, not all parameters were able to be estimated jointly in step 2. Therefore, in step 3, we fixed the reporting rate to its initially calibrated value and then re-fit the vaccine parameters, allowing for small changes that better captured the time trajectory in incidence. These initially estimated reporting rates optimized error for each age group, but did not minimize error for pooled cases across the age groups. Therefore, for the pooled population plots we hand adjusted the final reporting rate to match overall observed dynamics more closely. We note that because we assume the reporting rate is constant over time, estimates of vaccine efficacy and the trajectory of incidence over time are insensitive to the reporting rate used. An accurate reporting rate is only necessary to match incidence.

**Figure S3.** Comparison of estimates of vaccine efficacy over time for Sindh and Punjab (same material as Figure 2). Panels A) and C) show population impacts for Sindh (panel A) and Punjab (Panel C) with matching y-axis limits and panels B and D show age-specific impacts for Sindh (panel B) and Punjab (panel D).

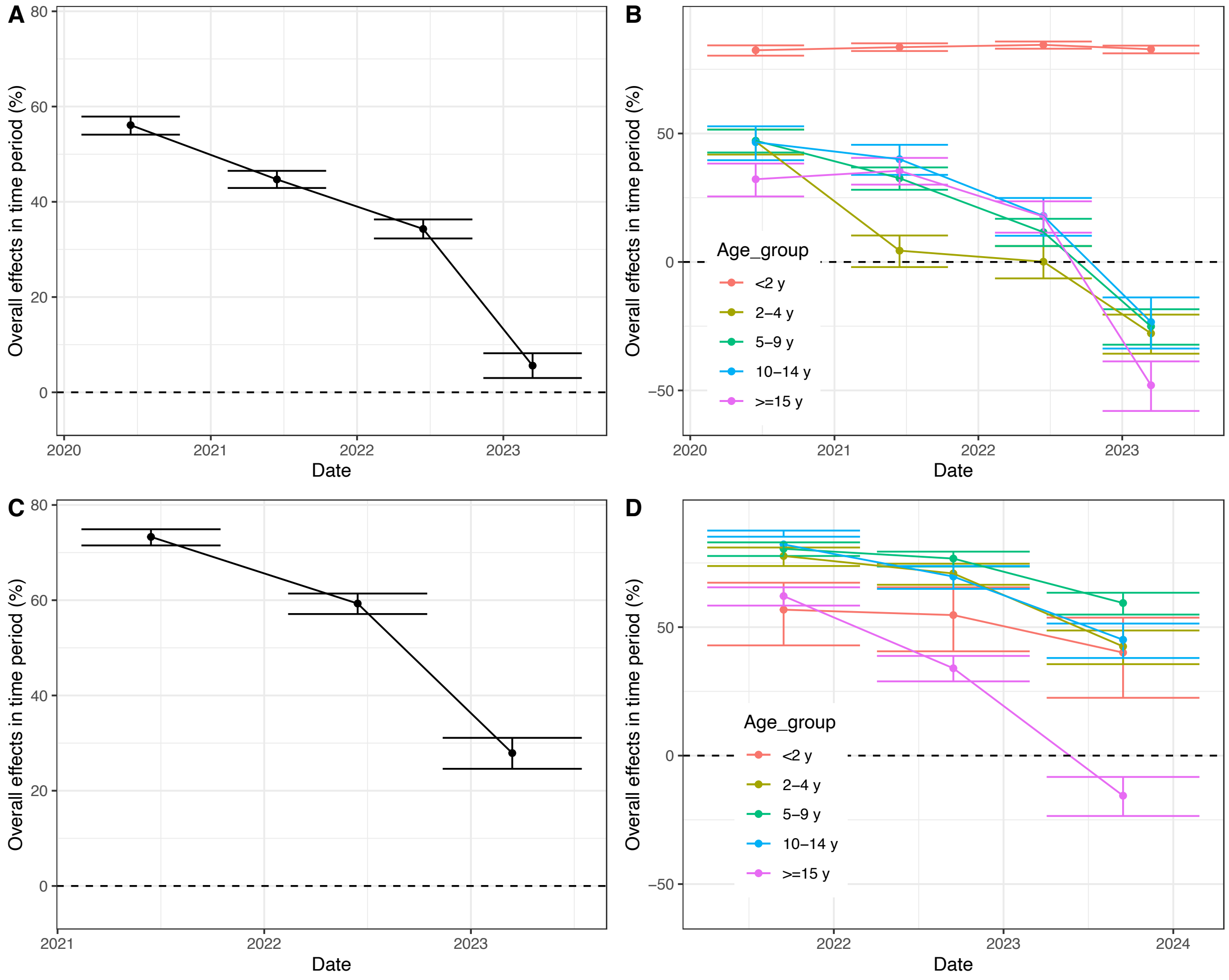

**Table S1.** Fixed model parameters constant across all contexts and simulations

| | Value |
|---|---|
| Typhoid carrier probability | 0.108 |
| Carrier removal year | 2500 |
| Relative infectiousness of chronic carriers | 0.241 |
| Environmental cutoff days | 20 |
| Environmental peak start | 275.85 |
| Environmental ramp down duration | 100 |
| Environmental ramp up duration | 175.26 |
| Prepatent relative infectiousness | 0.5 |
| Protection per infection | 0.98 |
| Subclinical relative infectiousness | 1 |
| Node Contagion Decay Rate | 0.24 |
| Life expectancy in 2017 | 66.51 years |

**Table S2.** Site-specific calibrated model parameters
*Parameter value is derived from the above parameters
**Computed as life expectancy (from table S1)/age of first infection

| | Sindh Fitted (credible range) | Punjab Fitted (credible range) |
|---|---|---|
| **Background transmission parameters** | | |
| Acute infectiousness | 42808 (0, 100000) | 9677 (0, 100000) |
| Exposure Lambda | 6.94 (-1, 15) | 2.36 (-1, 10) |
| Contact exposure rate (/day) | 0 (N/A) | 0 (N/A) |
| Environmental exposure rate (/day) | 1.99 (0, 4) | 0.61 (0, 4) |
| Symptomatic fraction (proportion) | 0.16 (0.01, 0.2) | 0.13 (0.01, 0.2) |
| **Vaccine parameters** | | |
| Duration of fixed immunity (days) | | |
| <2 y | 904.4 (0, 6935) | 0 (0, 6935) |
| 2-10 y | -- | 0 (0, 6935) |

| | | |
| --- | --- | --- |
| 12+ y | -- | 5448 (0, 6935) |
| 2-5 y | 240.9 (0, 6935) | -- |
| 5+ y | 0 (0, 6935) | -- |
| Duration of waning for immunity (days) | 505 (0, 6935) | 3278 (0, 6935) |
| Vaccine efficacy (proportion) | 0.97 (0.7, 1.0) | 1.0 (0.7, 1.0) |
| Routine immunization coverage (proportion) | 0.949 (0.25, 1) | 0.950 (0.1, 1) |
| Campaign coverage (proportion) | 0.660 (0.25, 1) | 0.995 (0.25, 1) |
| **Reporting rate (proportion)** | | |
| Age-specific (fitted) | 0.128 | 0.2 |
| Population (hand tuned) | 0.16 | 0.115 |
| **Average age of first infections (2015-2019)*** | 8.31 yrs | 11.99 yrs |
| **Basic reproduction number ($R_0$)**** | 8.00 | 5.55 |
| **Population growth rate** | 2.74%/year | 3.02%/year |

**Figure S4.** Age specific modeled (dashed) and data (solid) by age group for Sindh. Gray ribbons show 95% simulation intervals.

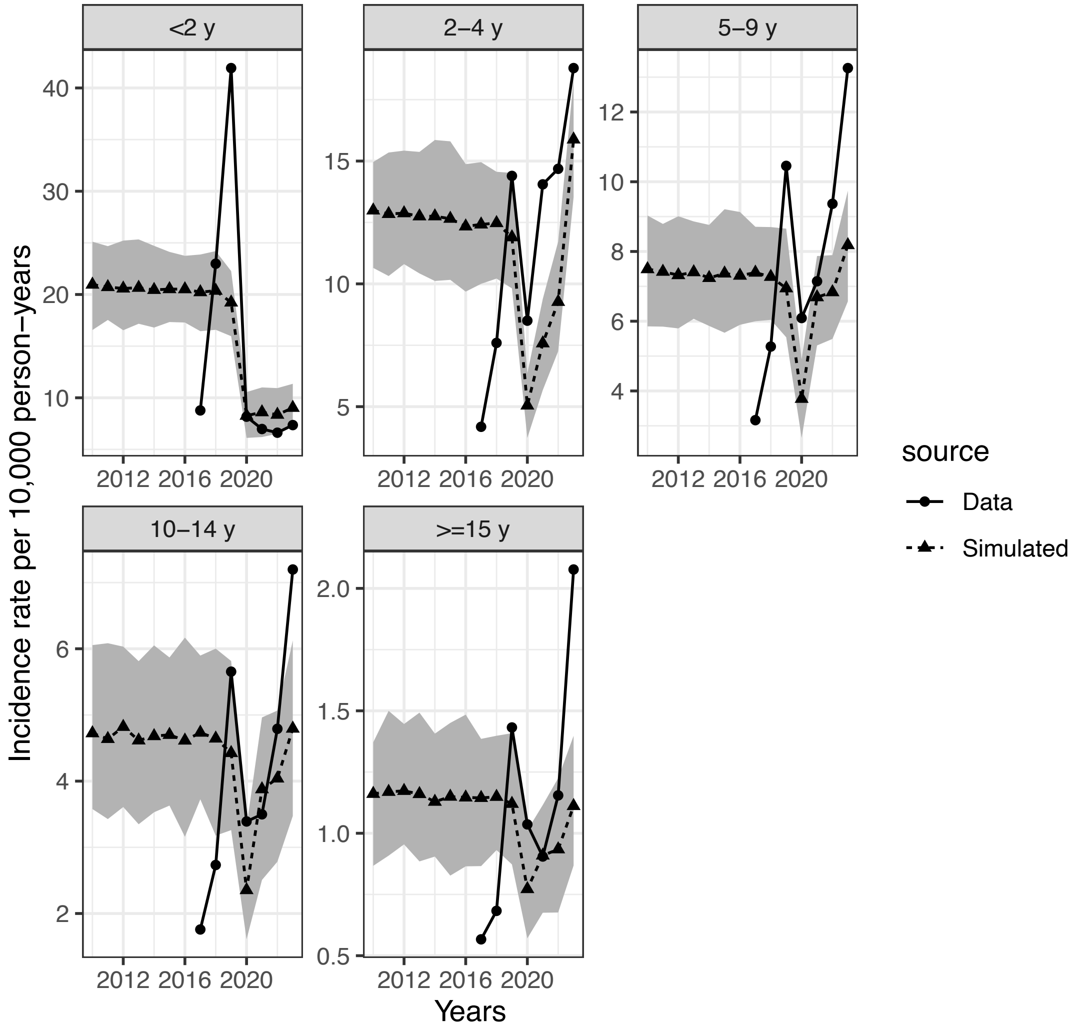

**Figure S5.** Age specific modeled (dashed) and data (solid) by age group period for Punjab. Gray ribbons show 95% uncertainty intervals.

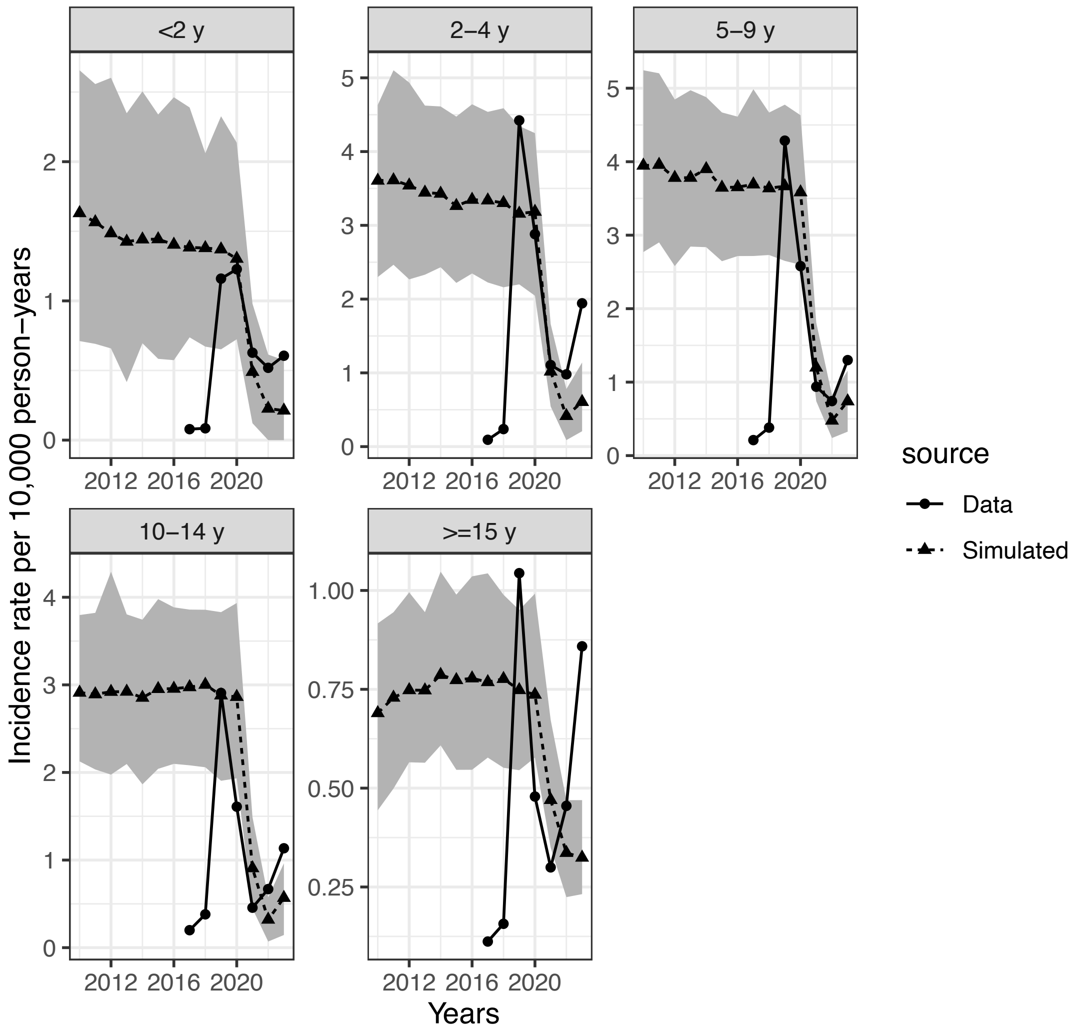

**Figure S6.** Alternative version of Figure 4A with 99.5% coverage for the forward vaccine campaign. Similar to main text simulations, routine immunization coverage among school children was modeled as being equal to campaign coverage, with coverage among adults modeled as being half as high as was attained among children under 15 years. Colors do not correspond to Figure 4A.

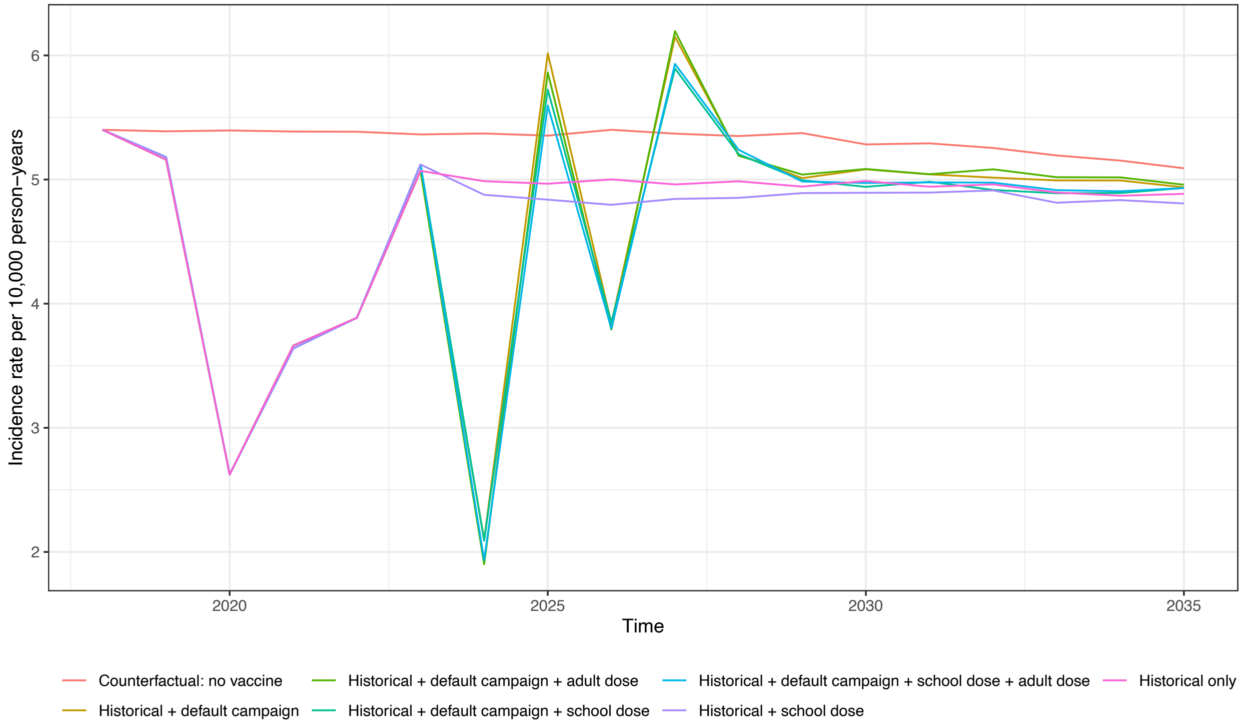
